# Infectious SARS-CoV-2 Virus in Symptomatic COVID-19 Outpatients: Host, Disease, and Viral Correlates

**DOI:** 10.1101/2021.05.28.21258011

**Authors:** Katie R. Mollan, Joseph J. Eron, Taylor J. Krajewski, Wendy Painter, Elizabeth R. Duke, Caryn G. Morse, Erin A. Goecker, Lakshmanane Premkumar, Cameron R. Wolfe, Laura J. Szewczyk, Paul L. Alabanza, Amy James Loftis, Emily J. Degli-Angeli, Ariane J. Brown, Joan A. Dragavon, John J. Won, Jessica Keys, Michael G. Hudgens, Lei Fang, David A. Wohl, Myron S. Cohen, Ralph S. Baric, Robert W. Coombs, Timothy P. Sheahan, William A. Fischer

## Abstract

**Background:** While SARS-CoV-2 infectious virus isolation in outpatients with COVID-19 has been associated with viral RNA levels and symptom duration, little is known about the host, disease and viral determinants of infectious virus detection.

**Methods:** COVID-19 adult outpatients were enrolled within 7 days of symptom onset. Clinical symptoms were recorded via patient diary. Nasopharyngeal swabs were collected to quantitate SARS-CoV-2 RNA by reverse transcriptase polymerase chain reaction and for infectious virus isolation in Vero E6-cells. SARS-CoV-2 antibodies were measured in serum using a validated ELISA assay.

**Results:** Among 204 participants with mild-to-moderate symptomatic COVID19, the median nasopharyngeal viral RNA was 6.5 (IQR 4.7-7.6 log_10_ copies/mL), and 26% had detectable SARS-CoV-2 antibodies (IgA, IgM, IgG, and/or total Ig) at baseline. Infectious virus was recovered in 7% of participants with SARS-CoV-2 antibodies compared to 58% of participants without antibodies (probability ratio (PR)=0.12, 95% CI: 0.04, 0.36; p=0.00016). Infectious virus isolation was also associated with higher levels of viral RNA (mean RNA difference +2.6 log_10_, 95% CI: 2.2, 3.0; p<0.0001) and fewer days since symptom onset (PR=0.79, 95% CI: 0.71, 0.88 per day; p<0.0001).

**Conclusions:** The presence of SARS-CoV-2 antibodies is strongly associated with clearance of infectious virus isolation. Seropositivity and viral RNA levels are likely more reliable markers of infectious virus clearance than subjective measure of COVID-19 symptom duration. Virus-targeted treatment and prevention strategies should be administered as early as possible and ideally before seroconversion.

**ClinicalTrials.gov Identifier:** NCT04405570

**Key Points (Summary):** Among COVID-19 outpatients within 7 days of symptom onset, the presence of SARS-CoV-2-specific antibodies was strongly associated with clearance of infectious virus. Seropositivity appears to be more reliable marker of infectious virus clearance than subjective measure of COVID-19 symptoms.

## Introduction

Severe acute respiratory syndrome coronavirus-2 (SARS-CoV-2), the etiological agent of coronavirus disease-2019 (COVID-19), has caused more than 175 million infections and 3.8 million deaths worldwide.^1,2^ The US Centers for Disease Control and Prevention (CDC) recommends physical isolation of 10 days for individuals with mild-to-moderate COVID-19, and 20 days for individuals with severe disease.^3^ However, infectious SARS-CoV-2 virus isolation has been reported beyond 20 days from patients with a compromised immune system.^4–9^ While prior studies have demonstrated an association between infectious virus isolation in the upper airway and high levels of nasal viral RNA measured by quantitative reverse transcription polymerase chain reaction (qRT-PCR),^10–14^ limited data are available regarding the host and disease factors associated with the presence of infectious virus in persons with mild-to-moderate COVID-19.

An improved understanding of viral and host factors associated with shedding of infectious virus is essential to prevent transmission of SARS-CoV-2 and reliably evaluate the antiviral efficacy of novel therapies. Here, we provide a comprehensive analysis of demographic, immunologic, virologic, and clinical disease factors associated with infectious virus isolation and levels of viral RNA in nasopharyngeal swab samples in the largest study of symptomatic outpatient adults with COVID-19.

## Methods

### Study design

This cross-sectional analysis was conducted at study entry among participants who enrolled in a Phase IIa clinical trial of the oral antiviral agent molnupiravir. Adult outpatients with symptomatic mild-to-moderate COVID-19 in the United States were enrolled within 1 week of symptom onset.^15,16^ The study protocol was approved by Western IRB (WIRB). Ten sites enrolled participants in: North Carolina, California, Washington, Texas, Florida, and Louisiana. All study participants provided written informed consent. Participants were eligible to enroll if they were ≥18 years of age, had SARS-CoV-2 infection with symptom onset within the past 7 days (168 hours), and were experiencing symptoms of COVID-19. Active SARS-CoV-2 infection was confirmed by molecular testing from a sample collected within 96hr prior to enrollment. A complete list of eligibility criteria is available at clinicaltrials.gov (NCT04405570).

### Measurement of SARS-CoV-2 RNA, Infectious Virus, and Antibodies

Nasopharyngeal (NP) swabs were collected from each participant at enrollment. Two NP swabs were collected—one from each nostril from all participants except for *n=*31 where a single NP swab was collected and aliquoted for testing. Nasopharyngeal swabs were placed into virus transport medium (VTM; supplied by COPAN), stored at 4°C prior to shipping in refrigerated containers to central laboratories, aliquoted, and stored at -80°C until tested. To measure viral RNA, qRT-PCR was performed on NP swabs using CDC primer/probe solution against the N1 region of the nucleocapsid gene (2019-nCoV_N1) at Covance CLS with Promega Maxwell for RNA extraction, manual plate build, and Quantstudio 12kflex for the 96 well qRT-PCR format (lower limit of quantification: 1,018 copies/mL).^17^

To measure the presence of infectious virus in each NP swab, samples were cultured in Vero E6 cells similar to prior studies,^10^ and this work was performed in a UNC Department of Environmental Health (EHS)/CDC-approved BSL3 facility following approved standard operating procedures and laboratory safety plans. Briefly, VTM samples were thawed, diluted 1:1 in infection medium and added in duplicate to Vero cell cultures plated 24hr prior. Quadruplicate negative (infection medium only) high (500 particle forming units, PFU) and low (50 PFU) positive controls were included.(11) On 2 and 5 days post inoculation (dpi), culture positivity was determined by measuring viral RNA in heat-inactivated culture medium by qRT-PCR using the Abbott m2000sp/rt quantitative SARS-CoV-2 RNA assay (limit of quantification: 100 copies/mL). The limit of culture detection was 50 PFU (MOI 0.0008) using two criteria for positivity: (i) SARS-CoV-2 RNA copies >1,000/mL in supernatant at 2dpi, or (ii) a relative change of >5 in RNA copy number from 2 to 5 dpi (i.e., [*copies_*5*dpi* / *copies*_2*dpi*] > 5).

SARS-CoV-2-specific antibodies in serum were analyzed using an antigen-capture enzyme-linked immunosorbent assay as described previously.^18^ Based on reference panel performance, the following optical density thresholds for seropositivity were applied: total Ig>0.376, IgG>0.376, IgA>0.30, IgM>0.31. SARS-CoV-2 antibody positive status was determined by a positive result on at least one of the following: total Ig, IgG, IgM, or IgA.

### Covariates

Participant demographics and disease characteristics were collected at baseline (study entry or screening), including medical history, hematology, and chemistry laboratory tests. Obesity was defined as body mass index (BMI) ≥30 kg/m^2^. Days since symptom onset was measured by participant report; and COVID-19 symptom severity was assessed using a four-point scale (absent, mild, moderate, or severe) in the participant-reported diary. Sixteen COVID-related symptoms were collected, as well as overall severity. D-dimer and C-reactive protein (CRP) were measured at local laboratories. D-dimer was dichotomized as <0.5 (within or near normal range) versus ≥0.5 mg/L, and CRP was dichotomized as <10 versus ≥10 mg/L.

### Data analyses

This analysis includes baseline results from enrolled participants. 95% confidence intervals (CIs) and p-values are presented with no adjustment for multiplicity. Missing data were excluded and typically were due to specimen cold-chain shipping abnormalities. Analyses were conducted in Windows SAS version 9.4 (Cary, NC) and Windows R version 4.0.2 or 4.0.4.

Associations between infectious virus isolation and each characteristic were estimated with a probability ratio (PR) and corresponding 95% CI from a modified Poisson model for bivariate and multivariable analyses.^19^ The following host characteristics were evaluated for associations with infectious virus isolation: presence of antibodies, symptom duration, age, sex, ethnicity (Hispanic versus non-Hispanic), BMI and obesity, frequent medical conditions known to be associated with worse COVID-19 illness (diabetes, hypertension, and asthma), overall symptom severity, and five COVID-19 respiratory-related symptoms (cough, shortness of breath, sore throat, nasal obstruction, nasal discharge). Race was not evaluated because few participants self-identified as African-American or Asian. Enrollment of Latinx participants varied considerably by site, and *post hoc* sensitivity analyses accounting for site were conducted using a Mantel-Haenszel pooled PR. The same statistical approach was used to estimate PRs for associations of each characteristic with seropositive antibody status.

Mean difference in SARS-CoV-2 RNA levels (log_10_ copies/mL) was estimated for each host characteristic listed above using a general linear model with heteroscedasticity-consistent standard errors. A receiver operator curve (ROC) analysis was applied to identify a SARS-CoV-2 RNA cut-point for infectious virus detection that minimized the Euclidean distance between the ROC curve and the (0,1) point in the ROC plane, to maximize sensitivity and specificity. Wilcoxon rank-sum tests were used to compare hematology measures by infectious virus status, and by antibody status. A chi-squared test was used to compare inflammatory markers (D-dimer, CRP) by infectious virus status, and by antibody status.

## Results

### Patient Population

Between 19 June 2020 and 22 January 2021, 240 participants were screened and 204 were enrolled. Enrollment visits occurred a median of 5 (IQR 4-5) days since symptom onset. Median age was 40 (IQR 27-52) years and 51% were women. Forty-seven percent of participants were White non-Hispanic, 42% Hispanic/Latinx, 5% Black non-Hispanic, 3% Asian non-Hispanic, and 2% multiple or other races (**Table 1**).

**Table 1.** Baseline characteristics of outpatients with mild-to-moderate COVID-19 (*n* = 204)

| Characteristic | Overall |
| --- | --- |
| **Sex** |  |
| Female | 104 (51%) |
| Male | 100 (49%) |
| **Age, years** |  |
| Median (Q1,Q3) | 40 (27, 52) |
| Min, Max | 18, 82 |
| **Race-ethnicity <sup>a</sup>** |  |
| Hispanic or Latinx, regardless of race | 86/203 (42%) |
| Two or more races, non-Hispanic | 3/203 (1%) |
| Asian, non-Hispanic | 6/203 (3%) |
| Black, non-Hispanic | 11/203 (5%) |
| White, non-Hispanic | 96/203 (47%) |
| Other, non-Hispanic | 1/203 (0%) |
| **Days since COVID-19 symptom onset** |  |
| Median (Q1,Q3) | 5 (4, 5) |
| Mean (SD) | 4.5 (1.4) |
| Min, Max | 0, 7 |
| **WHO ordinal scale <sup>b</sup>** |  |
| Ambulatory, limitation of activities | 151/194 (78%) |
| Ambulatory, no limitation of activities | 43/194 (22%) |
| **Body mass index <sup>a</sup>, kg/m<sup>2</sup>** |  |
| Median (Q1,Q3) | 27 (24, 31) |
| Min, Max | 18, 55 |
| Obesity: BMI ≥ 30 | 57/203 (28%) |
| **Frequent Medical Diagnoses** |  |
| Hypertension | 28/204 (14%) |
| Diabetes | 20/204 (10%) |
| Asthma | 19/204 (9%) |
| **Smoking/vaping status <sup>a, c</sup>** |  |
| Current | 30/203 (15%) |
| Former | 34/203 (17%) |
| Never | 139/203 (68%) |
| **Viral RNA from NP swab <sup>b</sup>, log<sub>10</sub> copies/mL** |  |
| Median (Q1,Q3) | 6.51 (4.68, 7.61) |
| Mean (SD) | 6.19 (1.84) |
| Min, Max | BLQ-9.93 |
| Below limit of quantification (<1,018 copies/mL) | 15/195 (8%) |
| **Infectious virus from NP swab <sup>b</sup>** |  |
| Positive, PCR(+) | 78/175 (45%) |
| Negative, PCR(-) | 97/175 (55%) |
| **SARS CoV-2 Antibodies <sup>b</sup>** |  |
| Seropositive on any: total Ig, IgM, IgG, or IgA | 46/177 (26%) |
| IgM(+) | 40/177 (23%) |
| IgG(+) | 36/177 (20%) |
| IgA(+) | 14/177 (8%) |
| **D-dimer <sup>b</sup>, mg/L FEU** |  |
| Median (Q1,Q3) | 0.28 (0.20, 0.44) |
| Min, Max | 0.15, 4.43 |
| <0.5 mg/L FEU (normal) | 146/183 (80%) |
| **C-Reactive Protein <sup>b</sup>, mg/L** |  |
| Median (Q1,Q3) | 3.0 (1.0, 8.0) |
| Min, Max | 0.3, 262.0 |
| <10 mg/L (normal) | 147/190 (77%) |
| **White blood cell count, x 10<sup>9</sup>/L** |  |
| Median (Q1,Q3) | 4.4 (3.7, 5.6) |
| Min, Max | 1.4, 18.3 |
| **Absolute lymphocyte count <sup>b</sup>, x 10<sup>9</sup>/L** |  |
| Median (Q1,Q3) | 1.4 (1.2, 1.9) |
| Min, Max | 0.5, 3.6 |
| **Absolute neutrophil count <sup>b</sup>, x 10<sup>9</sup>/L** |  |
| Median (Q1,Q3) | 2.4 (1.8, 3.3) |
| Min, Max | 0.4, 16.8 |
| **Absolute monocyte count <sup>b</sup>, x 10<sup>9</sup>/L** |  |
| Median (Q1,Q3) | 0.40 (0.30, 0.50) |
| Min, Max | 0.04, 1.10 |
| **Hemoglobin, g/dL** |  |
| Median (Q1,Q3) | 14.3 (13.4, 15.3) |
| Min, Max | 8.1, 17.7 |

<sup>a</sup> $n = 1$ missing. <sup>b</sup> Missing data: WHO ordinal scale ( $n = 10$ ), viral RNA ( $n = 9$ ), infectious virus ( $n = 29$ ), antibodies ( $n = 27$ ), D-dimer ( $n = 21$ ), C-reactive protein ( $n = 14$ ), absolute lymphocyte count ( $n = 34$ ), absolute neutrophil count ( $n = 34$ ), absolute monocyte count ( $n = 35$ ). <sup>c</sup> Smoking/vaping status is a composite of cigarette, marijuana, vaping, and cigar use.
BMI = body mass index; BLQ =below the limit of quantification ; NP = nasopharyngeal; PCR = Polymerase Chain Reaction; Q1 = 25<sup>th</sup> percentile; Q3 = 75<sup>th</sup> percentile; SARS = Severe acute respiratory syndrome; SD = standard deviation; US = United States; WHO = World Health Organization.

Baseline median SARS-CoV-2 viral RNA from NP swabs was 6.5 (IQR 4.7-7.6) log_10_ copies/mL and 8% of participants (15/195) had RNA below the limit of quantification (**Table 1**). Median viral RNA was highest within 3 days after symptom onset (**Supplementary Figure 1**). Overall symptom severity was rated as mild for 48%, moderate for 45%, and severe for 7% of participants. The most frequent symptoms, regardless of severity, were fatigue (80%), cough (77%), stuffy nose (74%), headache (71%), and muscle aches (64%). Loss of smell and taste were each experienced by 54% of participants (**Table 2**). Baseline laboratory testing was notable for a median baseline lymphocyte count of 1.4 ×10^9^/L (IQR 1.2-1.9). Most participants had D-dimer and CRP levels within or near the normal range with 80% below 0.5 mg/L and 77% below 10 mg/L, respectively (**Table 1**).

**Table 2.** Symptom severity among outpatients with mild-to-moderate COVID-19 ^a^.

| Symptom type | Severity <sup>b</sup>, n (row %) |  |  |  |
| --- | --- | --- | --- | --- |
|  | Any Severity | Mild | Moderate | Severe |
| ***Respiratory Symptoms (n = 202)*** |  |  |  |  |
| Cough | 155 (77%) | 95 (47%) | 48 (24%) | 12 (6%) |
| Nasal obstruction (stuffy nose) | 149 (74%) | 84 (42%) | 53 (26%) | 12 (6%) |
| Sore throat | 101 (50%) | 69 (34%) | 28 (14%) | 4 (2%) |
| Nasal discharge (runny nose) | 91 (45%) | 64 (32%) | 22 (11%) | 5 (2%) |
| Shortness of breath | 83 (41%) | 56 (28%) | 22 (11%) | 5 (2%) |
| ***Other Symptoms (n = 202)*** |  |  |  |  |
| Fatigue | 162 (80%) | 73 (36%) | 59 (29%) | 30 (15%) |
| Headache | 144 (71%) | 70 (35%) | 52 (26%) | 22 (11%) |
| Muscle aches | 130 (64%) | 62 (31%) | 54 (27%) | 14 (7%) |
| Loss of smell | 109 (54%) | 23 (11%) | 34 (17%) | 52 (26%) |
| Loss of taste | 109 (54%) | 28 (14%) | 35 (17%) | 46 (23%) |
| Chills | 83 (41%) | 54 (27%) | 20 (10%) | 9 (4%) |
| Feeling feverish | 82 (41%) | 54 (27%) | 22 (11%) | 6 (3%) |
| Diarrhea | 64 (32%) | 43 (21%) | 18 (9%) | 3 (1%) |
| Nausea | 57 (28%) | 34 (17%) | 16 (8%) | 7 (3%) |
| Fainting | 15 (7%) | 13 (6%) | 1 (<1%) | 1 (<1%) |
| Vomiting | 14 (7%) | 6 (3%) | 6 (3%) | 2 (1%) |
| ***Overall Symptom Severity (n = 197)*** |  | 95 (48%) | 89 (45%) | 13 (7%) |

<sup>a</sup> *n* = 2 participants were missing a symptom diary, and *n* = 5 additional participants did not report Overall Symptom Severity. <sup>b</sup> Participants ranked each reported symptom as Absent, Mild, Moderate, or Severe using a symptom diary. The ‘Any Severity’ column is a sum of Mild, Moderate, or Severe (i.e., regardless of severity level).

### SARS-CoV-2 antibodies

Of 177 patients with evaluable measurements, 26% were antibody positive at baseline for at least one SARS-CoV-2 spike protein-specific immunoglobulin isotype, with 20% IgG+, 23% IgM+, and 8% IgA+ (**Table 1**). Three participants were IgG(+) with IgM(-) and IgA(-) and one was IgA+ only (**Supplementary Table 1**). In bivariate analyses, host and disease characteristics associated with having SARS-CoV-2 specific antibodies included longer time since symptom onset (probability ratio [PR]=1.27 per 1 day, 95%CI: 1.06, 1.52, p=0.0091) and higher white blood cell counts, including total white blood cells and absolute neutrophil count (**Supplementary Figure 2**). Participants with SARS-CoV-2 antibodies were more likely to have elevated D-dimer and CRP levels as compared to seronegative participants (D-dimer ≥0.5 mg/L: 39% versus 13%, p=0.00037; CRP ≥10 mg/L: 44% versus 15%, p=0.00011; **Supplementary Table 2**).

### Correlates of virus isolation

Infectious virus was isolated from 45% (78/175) of participants at baseline. Those with positive virus isolation had a median symptom duration of 4 (IQR 3-5) days compared to 5 (IQR 4-6) days in participants with negative virus isolation. Mean viral RNA levels were higher among participants with infectious virus isolation compared to those who were culture negative (mean: 7.6 versus 5.0 log_10_ copies/mL; mean difference: 2.6, 95%CI: 2.2, 3.0, p<0.0001; **Figure 1**). An ROC curve analysis revealed that a viral RNA threshold of ≥6.4 log_10_ copies/mL was predictive of infectious virus isolation (**Supplementary Figure 3**), with an estimated positive predictive value of 80% and negative predictive value of 91% (**Supplementary Table 3**).

**Figure 1.**
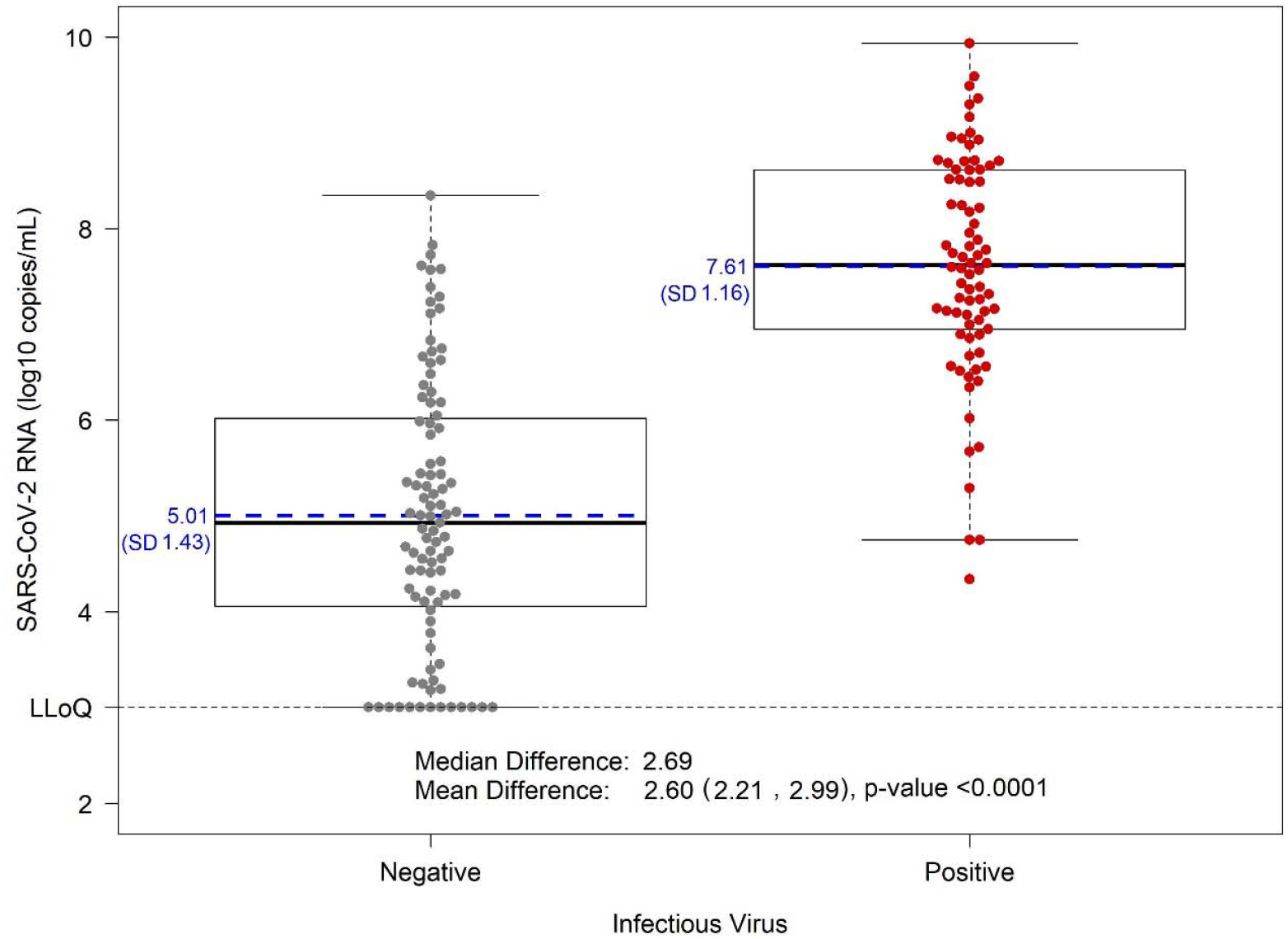
SARS-CoV-2 viral RNA levels in nasopharyngeal swab by infectious virus status. Nasopharyngeal viral RNA levels measured via qRT-PCR are displayed by infectious virus status, with culture negative in grey (*n* = 95) and culture positive (*n* = 78) in red; each dot represents a participant. Solid lines on the boxplots display the median and 25th-75th percentile (mean = blue dashed line) and the whiskers extend to the extrema (no more than 1.5 times the IQR from the box). LLoQ = lower limit of quantification; SD = standard deviation.

Infectious virus was isolated from 7% (3/43) of seropositive compared to 58% (73/125) of seronegative participants, consistent with an 88% lower prevalence of infectious virus for those with SARS-CoV-2 antibodies (probability ratio [PR]=0.12, 95%CI: 0.04, 0.36, p=0.00016). Infectious virus was isolated in only three seropositive individuals at 3, 4, and 6 days from symptom onset (all 3 had SARS-CoV-2 specific IgM; 2 had IgG; and 0 had IgA). There was also a strong association between antibody status and NP viral RNA levels (mean: 4.4 log_10_ copies/ml among seropositive versus 6.8 log_10_ copies/ml among seronegative participants; mean difference: -2.4, 95%CI: -2.8, -1.9, p<0.0001). Those with viral RNA levels in the top quartile (>7.6 log_10_) were seronegative (100%; 43/43) with the majority (91%; 39/43) having infectious virus detected, whereas those in the bottom quartile (<4.7 log_10_) were almost entirely infectious virus negative (98%; 40/41) and most were seropositive (68%; 28/41) (**Figure 2**).

**Figure 2.**
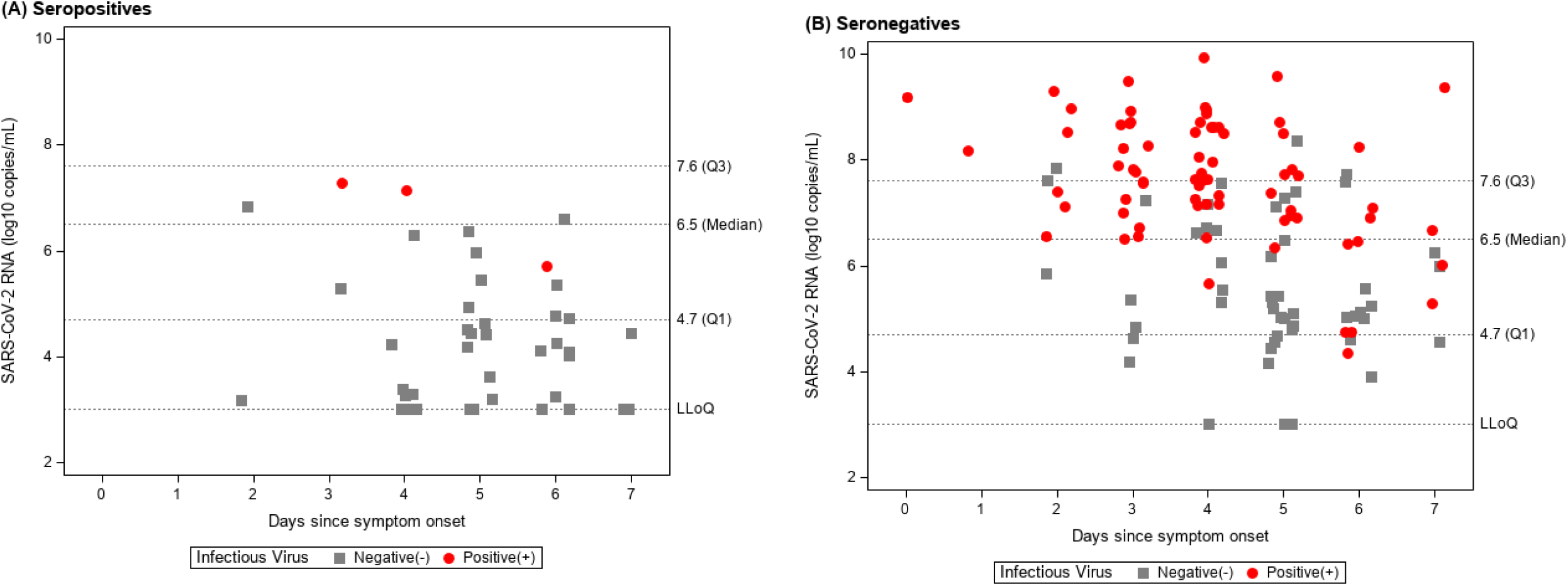
SARS-CoV-2 viral RNA, time since symptom onset, and infectious virus by SARS-CoV-2 specific antibody serostatus. Panel A shows seropositive participants. Panel B shows seronegative participants. Seropositive is defined as having SARS-CoV-2 specific total Ig, IgG, IgM, or IgA antibodies. Nasopharyngeal viral RNA levels (log_10_ copies/mL) are shown on the y-axis and days since symptom onset on the x-axis, with infectious virus culture positive participants in red circles, and culture negative participants in grey squares. The overall viral RNA median (Q1, Q3) and LLoQ are indicated by dashed horizontal lines. Ig = immunoglobulin; LLoQ = lower limit of quantification; Q1 = 25^th^ percentile; Q3 = 75^th^ percentile.

In a multivariable model (including viral RNA, antibody status, and symptom duration), infectious virus isolation was more prevalent for those with higher viral RNA (PR=1.52 per 1.0 log_10_ copies/mL, 95%CI: 1.37, 1.70, p<0.0001) and less prevalent for those with antibodies (PR=0.35 for seropositive versus seronegative, 95%CI: 0.11, 1.07, p=0.066), but was no longer associated with days since symptom onset (PR=1.01 per 1 day, 95%CI: 0.91, 1.11, p=0.90).

To evaluate the association between infectious virus isolation and host characteristics both bivariate and multivariable analyses were performed. In bivariate analyses, infectious virus isolation was more prevalent among participants with hypertension (PR=1.62, 95%CI: 1.15, 2.28, p=0.0054) and participants with nasal obstruction (PR=1.53, 95%CI: 0.97, 2.41, p=0.066). However, infectious virus isolation was less prevalent among participants who were further from symptom onset (PR=0.79 per 1 day, 95%CI: 0.71, 0.88, p<0.0001), those with moderate-to-severe versus mild symptoms (PR=0.68, 95%CI: 0.48, 0.96, p=0.029), those with shortness of breath (PR=0.52, 95%CI: 0.35, 0.77, p=0.0013), and those who self-identified as Latinx compared to non-Latinx (PR=0.54, 95%CI: 0.36, 0.79, p=0.0016). In a sensitivity analysis adjusting for site, the association between Latinx ethnicity and lower prevalence of virus isolation remained (Mantel-Haenszel PR=0.52, 95%CI: 0.28, 0.96, p=0.032). In this study, infectious virus isolation was not clearly associated with age (PR=1.09 per 10 years, 95%CI: 0.98, 1.22, p=0.10) or biological sex (PR=0.85 women vs. men, 95%CI: 0.61, 1.19, p=0.34) (**Figure 3**). In a multivariable model including host characteristics (**Supplementary Table 4**), seronegativity, fewer days since symptom onset, non-Latinx ethnicity, and hypertension remained associated with higher prevalence of infectious virus isolation.

**Figure 3.**
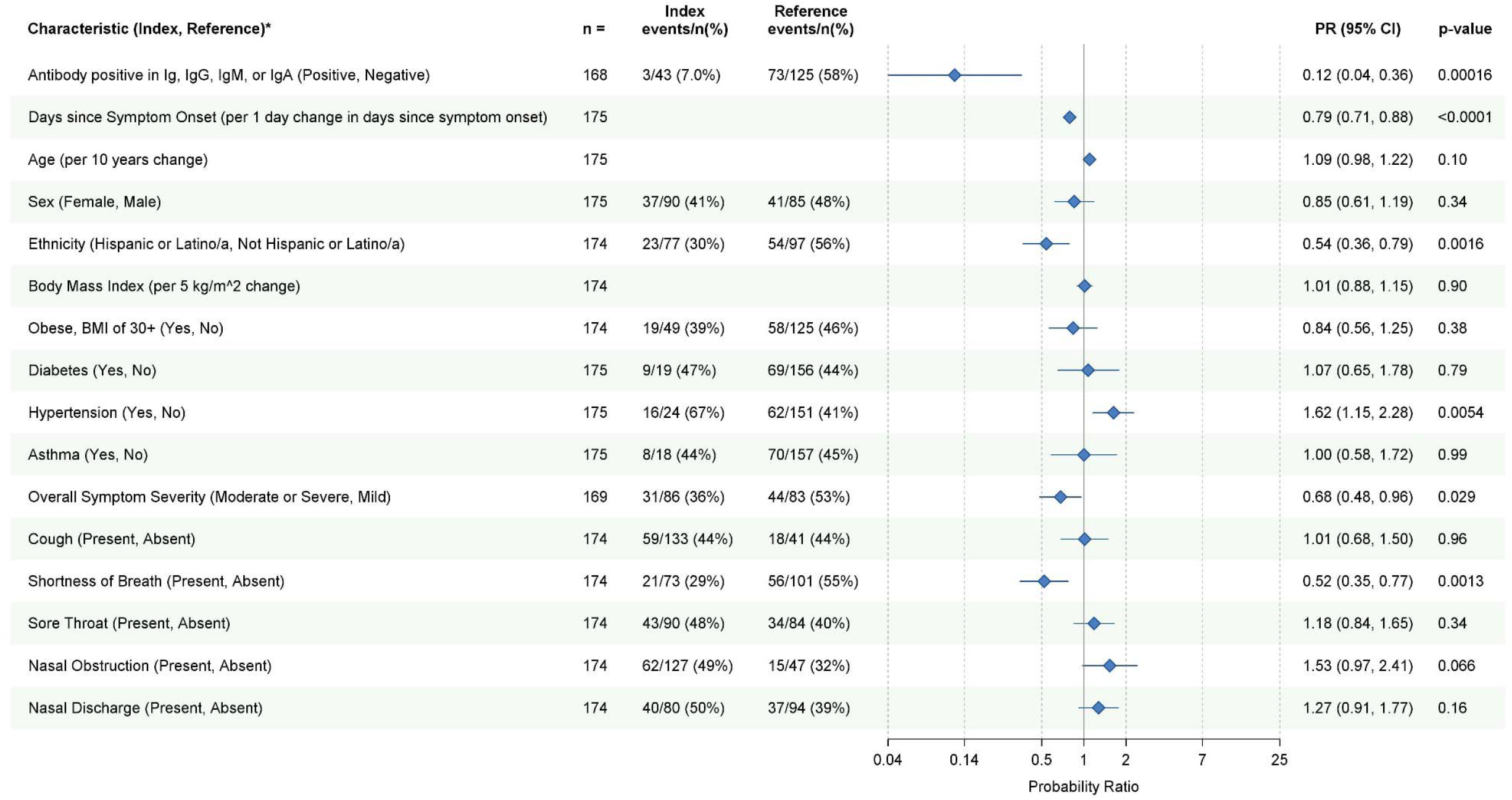
Host factor associations with infectious SARS-CoV-2 virus isolation. Bivariate unadjusted analyses are shown. *Prevalence ratios for the probability of infectious virus isolation were estimated for each dichotomous characteristic and a prevalence ratio per unit change was estimated for each continuous characteristic with corresponding 95% confidence intervals. Each continuous characteristic was fit as linear in the log-prevalence of infectious virus isolation. PR = prevalence ratio.

Bivariate associations between infectious virus isolation and inflammatory and hematology markers were also evaluated. Elevated inflammatory markers were numerically less frequent among participants with infectious virus culture positive compared to culture negative (**Supplementary Table 2**). Participants with positive infectious virus culture had lower levels of white blood cells including total white blood cell count, absolute lymphocyte count, and absolute neutrophil count **(Supplementary Figure 4)**.

### Correlates of viral RNA

Bivariate analyses of host factor associations with viral RNA level are provided in **Supplementary Figure 5**. Higher viral RNA was associated with fewer days since symptom onset (mean difference (MD) -0.5 log_10_ per 1 day, 95%CI: -0.6, -0.3, p<0.0001). There was a trend toward higher viral RNA levels in those with mild symptom severity (MD: -0.5 log_10_ for moderate-to-severe versus mild symptoms, 95%CI: -1.0, 0.07, p=0.086), with median viral RNA levels of 6.8 (IQR 4.8-8.1), 6.4 (IQR 4.6-7.3), and 5.7 (IQR 4.6-7.1) log_10_ copies/mL for participants with mild, moderate, and severe overall symptom severity, respectively. Viral RNA levels were higher on average for those with hypertension (MD: 1.0 log_10_, 95%CI: 0.3, 1.7, p=0.0036) or nasal discharge (MD: 0.7 log_10_, 95%CI: 0.2, 1.2, p=0.0052). Lower viral RNA levels were observed for those who self-identified as Latinx; however, in a sensitivity model adjusting for site as a covariate, the association between Latinx ethnicity and viral RNA was no longer present (MD: -0.01 log_10_, 95%CI: -0.7, 0.7, p=0.98). Viral RNA was not associated with age (MD: 0.1 log_10_ per 10 years of age, 95%CI: -0.06, 0.3, p=0.20) or body mass index (MD: 0.02 log_10_ per 5 kg/m^2^, 95%CI: -0.2, 0.3, p=0.83). There was some evidence of lower levels of viral RNA in women compared to men (MD: -0.5 log_10_, 95%CI: -1.0, 0.02, p=0.057).

## Discussion

SARS-CoV-2 infectious virus isolation within 1 week of symptom onset was prevalent and strongly associated with both viral RNA levels and the absence of SARS-CoV-2-specific antibodies, suggesting that humoral immunity plays a key role in limiting infectious virus shedding. This observation is supported by studies identifying prolonged viral replication in individuals with immune deficiencies, nearly all of whom had specific deficiencies in B cell function.^4–8,20^ Importantly, the novel association, in this study, of seropositivity with clearance of infectious virus supports recent reports of increased clinical efficacy of monoclonal antibodies in seronegative patients with COVID-19,^21^ and indicates that seropositivity could be a more reliable indicator of time since infection than duration of symptoms and could serve to identify those who would benefit the most from antiviral therapy.

At entry, most individuals in this study had a cough, nasal obstruction, fatigue, or headache; about half reported sore throat or loss of taste/smell; and 41% reported feeling feverish, having chills, or experiencing shortness of breath. Our study revealed associations between infectious virus isolation and milder symptoms, an absence of shortness of breath, and lower white blood cell populations, consistent with an early stage in the clinical course of infection. Presence of SARS-CoV-2 antibodies was associated with higher white blood cell and neutrophil counts, which is also consistent with these findings. Prior studies report no virus isolation beyond 10 days from symptom onset in outpatients with mild-moderate COVID-19.^11,14,22,23^ In this study, when accounting for both NP RNA level and serologic response, duration of symptoms (up to 7 days) was no longer associated with infectious virus isolation.

Viral RNA was highest soon after symptom onset with a peak in the median levels ≤ 3 days after symptom onset, and lower levels for those presenting 5 to 7 days after symptom onset. In this study, viral RNA in NP swabs of 6.4 log_10_ copies/mL or higher was predictive of infectious virus detection, with a positive predictive value of 80% and negative predictive value of 91%, supporting prior studies suggesting a threshold of 6 log_10_ copies/mL.^10,11,24^ However, infectious virus was isolated in three individuals with viral RNA below 5.0 log10 copies/mL (minimum: 4.3 log10 copies/mL), all three of whom were antibody negative men at 6 days from symptom onset, suggesting a delayed immune response.

We did not identify associations between several established risk factors for severe COVID-19 –including age, obesity, diabetes or asthma– and infectious virus isolation or higher levels of viral RNA. However, individuals with hypertension did tend to have higher levels of viral RNA and higher prevalence of infectious virus isolation, the mechanism of which remains unclear. Importantly, most outpatients with mild-to-moderate symptoms, even those in higher risk groups, have a relatively low probability of developing severe COVID-19 disease.^25–27^

This study describes novel associations between virologic and immunologic factors and isolation of infectious virus, however there are important limitations to consider. This was a cross-sectional sample of predominantly low risk, symptomatic outpatients, restricting our target population to those with mild-to-moderate COVID-19. The inclusion of only symptomatic participants precludes an assessment of factors associated with infectious virus among asymptomatic individuals. This study enrolled a large number of participants who identified as Latinx, yet there was an under-representation of African-Americans and Asian-Americans compared with the US burden of COVID-19, limiting generalizability.^28^ Further, seropositivity is not synonymous with virus neutralization. Infectious virus was isolated from three seropositive participants, two of whom also had RNA levels above 6.0 log_10_ but neutralization titers were not measured, thereby limiting this analysis to a qualitative assessment of seropositivity rather than antibody function. We also acknowledge that a negative culture does not entirely exclude the presence of infectious virus. The sensitivity of the virus isolation assay is 50pfu/ml, and thus infectious virus below this limit may not be detected. Additionally, variability in swabbing technique can result in false negative cultures.

Nonetheless, the strong associations we observed between antibody status and viral RNA shedding with isolation of infectious virus provide important insight into viral and host factors associated with infectious SARS-CoV-2 virus. Measuring a patient’s serostatus and NP RNA level can help clinicians assess the likelihood of infectious virus detection, which can help guide isolation practices, therapy, and clinical follow-up. Longitudinal research is needed to evaluate whether the early detection of SARS-CoV-2 antibodies can predict subsequent infectious virus clearance and whether early viral clearance decreases the likelihood of long-term COVID-19 symptoms.^29^ As SARS-CoV-2 continues to spread globally, improved understanding of host, disease, and viral factors associated with infectious virus isolation is important to inform non-pharmacologic, vaccine, and therapeutic strategies to interrupt transmission and progression of disease.

## Data Availability

Upon approval from the corresponding author and Ridgeback Biotherapeutics, in collaboration with Merck & Co., de-identified participant level data may be shared given the investigator who requests the data has approval from an Institutional Review Board, Independent Ethics Committee, or Research Ethics Board, as applicable, and executes a data use/sharing agreement.

## Acknowledgements

We thank Dr. Sarah Reifeis for data carpentry support, Ms. Ann Marie Weideman for statistical graphics advice, and Covance by Labcorp for data management support and quantification of viral RNA. We sincerely thank all the participants and sites for their dedication and contributions to the study: Valley Clinical Trials (Northridge, CA), University of North Carolina (Chapel Hill, NC), Fred Hutchinson (Seattle, WA), Care United Research (Forney, TX), Benchmark Research (Colton, CA), FOMAT Medical Research (Oxnard, CA), Indago Research & Health Center (Hiahleah, FL), Wake Forest Baptist Health (Winston-Salem, NC), Duke University (Durham, NC), and NOLA Research Works (New Orleans, LA). Molnupiravir was invented at Drug Innovations at Emory (DRIVE) LLC, a not-for-profit biotechnology company wholly owned by Emory University and is being developed by Merck & Co., Inc. in a collaboration with Ridgeback Biotherapeutics.

## Author contributions

KRM, JJE, TPS, and WAF led the conceptualization of the analysis and the writing of original manuscript draft. EAG, LP, EJDA, AJB, JAD, JJW, RSB, RWC, and TPS conducted SARS-CoV-2 laboratory assessments. KRM, TJK, JK, and MGH conducted the statistical analyses. Study data were verified by KRM, TJK, WP, LJS, LF, and WAF. KRM, JJE, WP, LJS, RSB, MGH, LF, DAW, RSB, TPS, and WAF contributed to the study design. WP, ERD, CGM, CRW, LJS, PLA, AJL, and WAF oversaw data collection or protocol implementation. All authors reviewed the draft manuscript and approved the final version.

---

**Supplementary Table 1.** SARS-CoV-2 specific antibody combinations.

| Measure | Events/n (%) |
| --- | --- |
| IgG(-), IgM(-), IgA(-) | 133/177 (75%) |
| IgG(+), IgM(+), IgA(-) | 20/177 (11%) |
| IgG(+), IgM(+), IgA(+) | 13/177 (7%) |
| IgG(-), IgM(+), IgA(-) | 7/177 (4%) |
| IgG(+), IgM(-), IgA(-) | 3/177 (2%) |
| IgG(-), IgM(-), IgA(+) | 1/177 (1%) |

CoV = coronavirus; Ig = immunoglobulin; SARS = Severe acute respiratory syndrome.

**Supplementary Table 2.** Associations between SARS-CoV-2 measures and inflammatory markers.

| SARS-CoV-2 Measure | Group | D-dimer (mg/L) |  | p-value <sup>a</sup> | C-Reactive Protein (mg/L) |  | p-value <sup>a</sup> |
| --- | --- | --- | --- | --- | --- | --- | --- |
|  |  | Below 0.5<br>(normal) | 0.5 or higher<br>(elevated) |  | Below 10<br>(normal) | 10 or above<br>(elevated) |  |
| Infectious virus | Culture positive | 58/69 (84%) | 11/69 (16%) | 0.21 | 59/71 (83%) | 12/71 (17%) | 0.12 |
|  | Culture negative | 66/87 (76%) | 21/87 (24%) |  | 67/92 (73%) | 25/92 (27%) |  |
| Antibodies | Seropositive | 25/41 (61%) | 16/41 (39%) | 0.00037 | 24/43 (56%) | 19/43 (44%) | 0.00011 |
|  | Seronegative | 104/120 (87%) | 16/120 (13%) |  | 104/123 (85%) | 19/123 (15%) |  |

<sup>a</sup> Chi-square p-value
CoV = coronavirus; SARS = Severe acute respiratory syndrome.

**Supplementary Table 3.** Nasopharyngeal viral RNA threshold for infectious virus isolation.

| **Viral RNA** | **Infectious Virus** |  |  |
| --- | --- | --- | --- |
| No. of participants<br>Row %<br>Column % |  |  |  |
|  | **Culture Negative** | **Culture Positive** | **Total** |
| **< 6.4 log<sub>10</sub> copies/mL** | 77<br>NPV: 91%<br>Sp: 81% | 8<br>9%<br>10% | 85 |
| **≥ 6.4 log<sub>10</sub> copies/mL** | 18<br>20%<br>19% | 70<br>PPV: 80%<br>Se: 90% | 88 |
| **Total** | 95 | 78 | 173 |

NPV = negative predictive value; PPV = positive predictive value; Se = sensitivity; Sp = specificity.

**Supplementary Table 4.** Multivariable model of host factor associations with infectious SARS-CoV-2 virus isolation (*n* = 162) ^a^.

| Characteristic (Index, Reference) | Adjusted Prevalence Ratio | 95% Confidence Interval | p-value |
| --- | --- | --- | --- |
| Antibody positive in Ig, IgG, IgM, or IgA (Positive, Negative) | 0.15 | (0.05, 0.47) | 0.0011 |
| Symptom Onset (per 1 day change in days since symptom onset) | 0.86 | (0.76, 0.97) | 0.016 |
| Age (per 10 years change) | 1.06 | (0.95, 1.18) | 0.32 |
| Sex (Female, Male) | 0.82 | (0.61, 1.11) | 0.20 |
| Ethnicity (Hispanic or Latinx, Not Hispanic or Latinx) | 0.63 | (0.45, 0.90) | 0.011 |
| Obese, BMI of 30+ (Yes, No) | 0.83 | (0.58, 1.21) | 0.34 |
| Diabetes (Yes, No) | 0.84 | (0.48, 1.45) | 0.52 |
| Hypertension (Yes, No) | 1.57 | (1.13, 2.19) | 0.0074 |
| Asthma (Yes, No) | 0.96 | (0.58, 1.60) | 0.88 |
| Overall Symptom Severity (Moderate-to-severe, Mild) | 0.97 | (0.70, 1.33) | 0.85 |

BMI = body mass index; Ig = immunoglobulin.
<sup>a</sup> Host characteristics in the table were included in this multivariable (modified Poisson) model to estimate adjusted prevalence ratios for the outcome of infectious virus isolation (i.e., culture positive). A prevalence ratio per unit change was estimated for each continuous characteristic, with days since symptom onset and age each fit as linear in the log-prevalence of infectious virus isolation. Participants with missing data were excluded.

## Figures

**Supplementary Figure 1.**
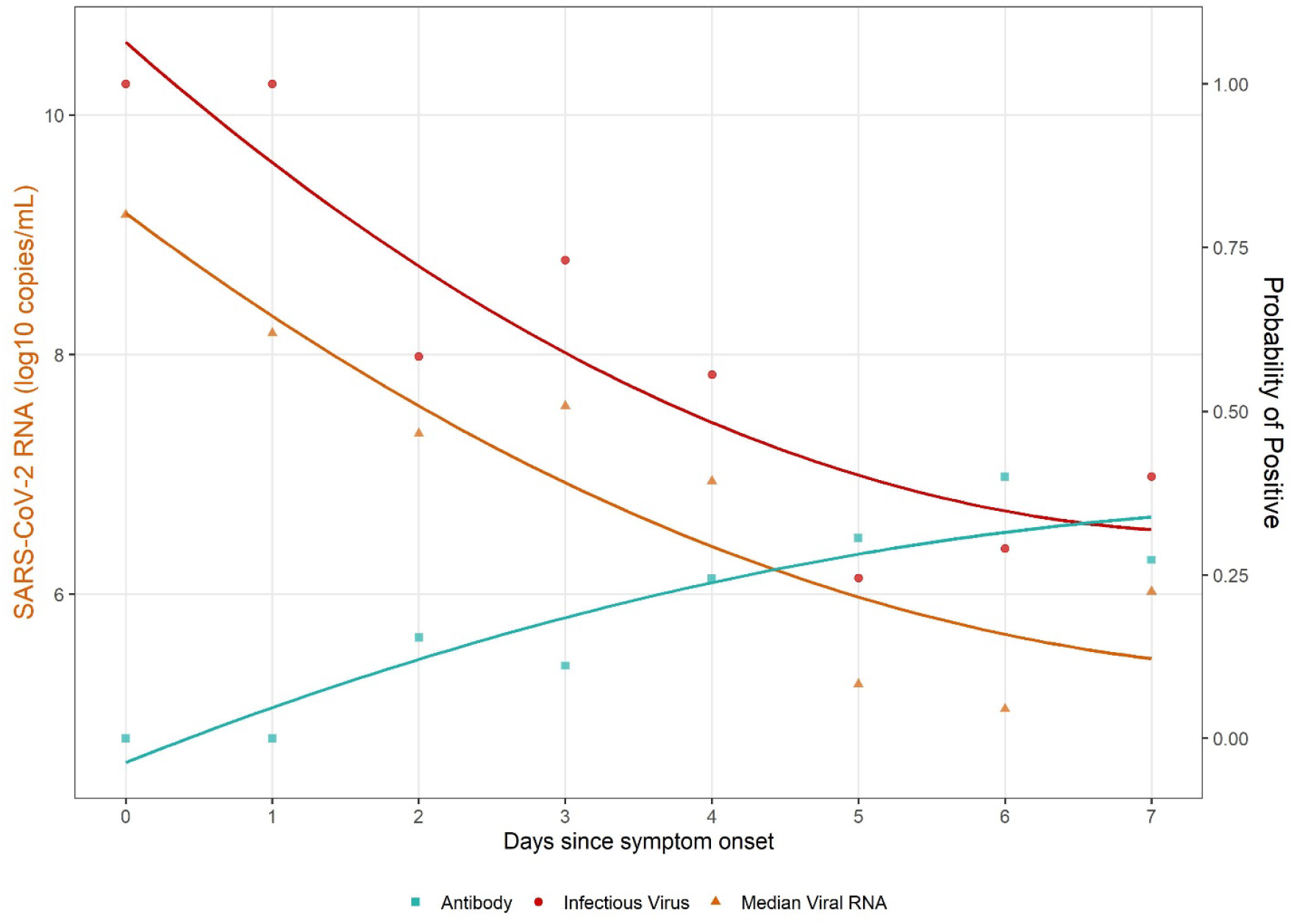
SARS-CoV-2 virus isolation, viral RNA, and serostatus according to days since symptom onset. Estimated prevalence of antibody positive (teal), prevalence of infectious virus positive (red), and the median viral RNA level (orange) are shown according to day since symptom onset. The left y-axis corresponds to viral RNA level and the right y-axis shows both the prevalence of infectious virus and prevalence of antibody positivity. Trend lines were estimated using a quadratic fit.

**Supplementary Figure 2.**
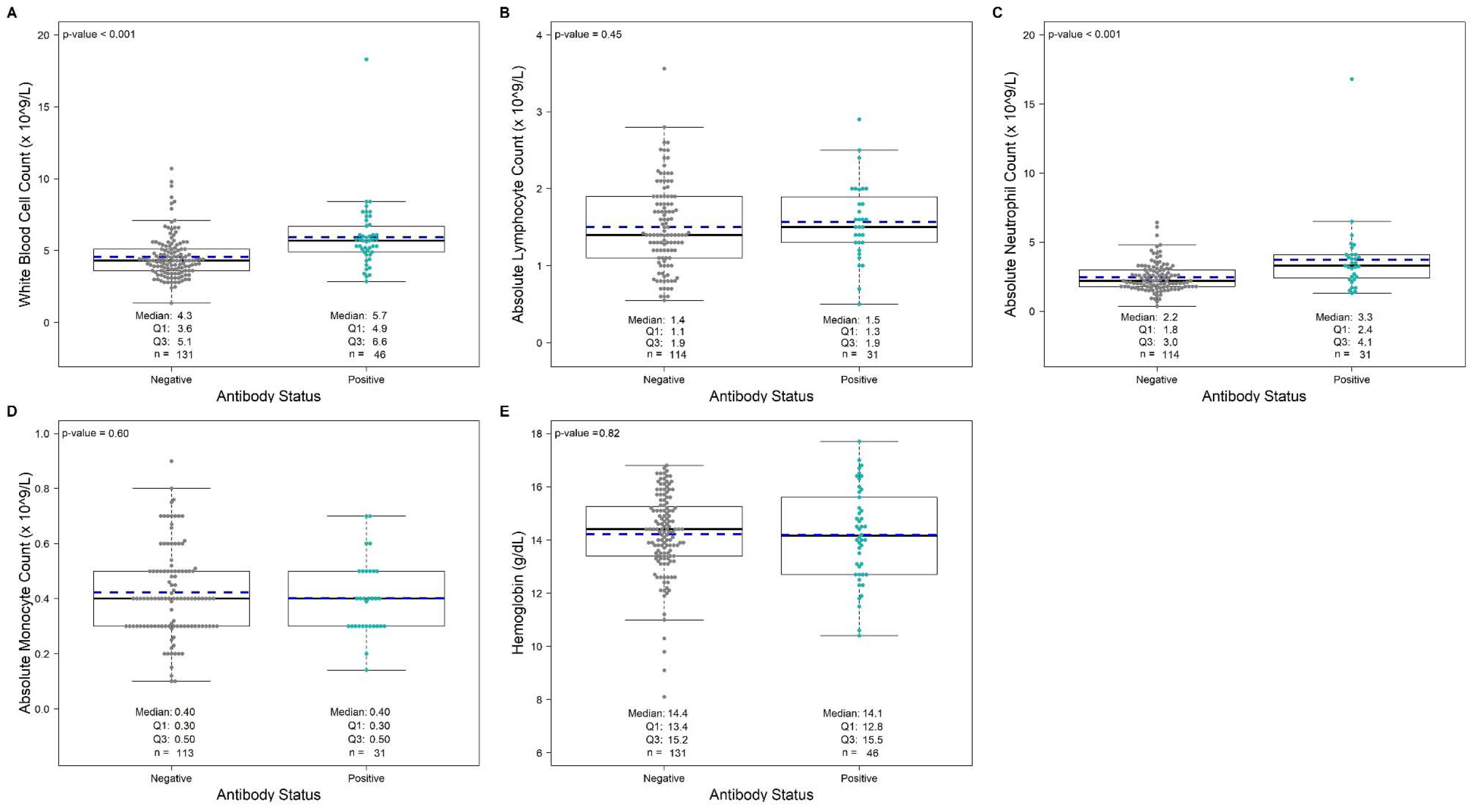
Associations between hematology measures and SARS-CoV-2 specific antibody status. Panels A-E: Five hematology measures are displayed by SARS-CoV-2-specific antibody status (total Ig, IgG, IgM, or IgA positivity), with antibody negative shown in grey and antibody positive shown in teal; each dot represents a participant. Solid lines on boxplots display the median and 25th-75th percentile (mean = blue dashed line).

**Supplementary Figure 3.**
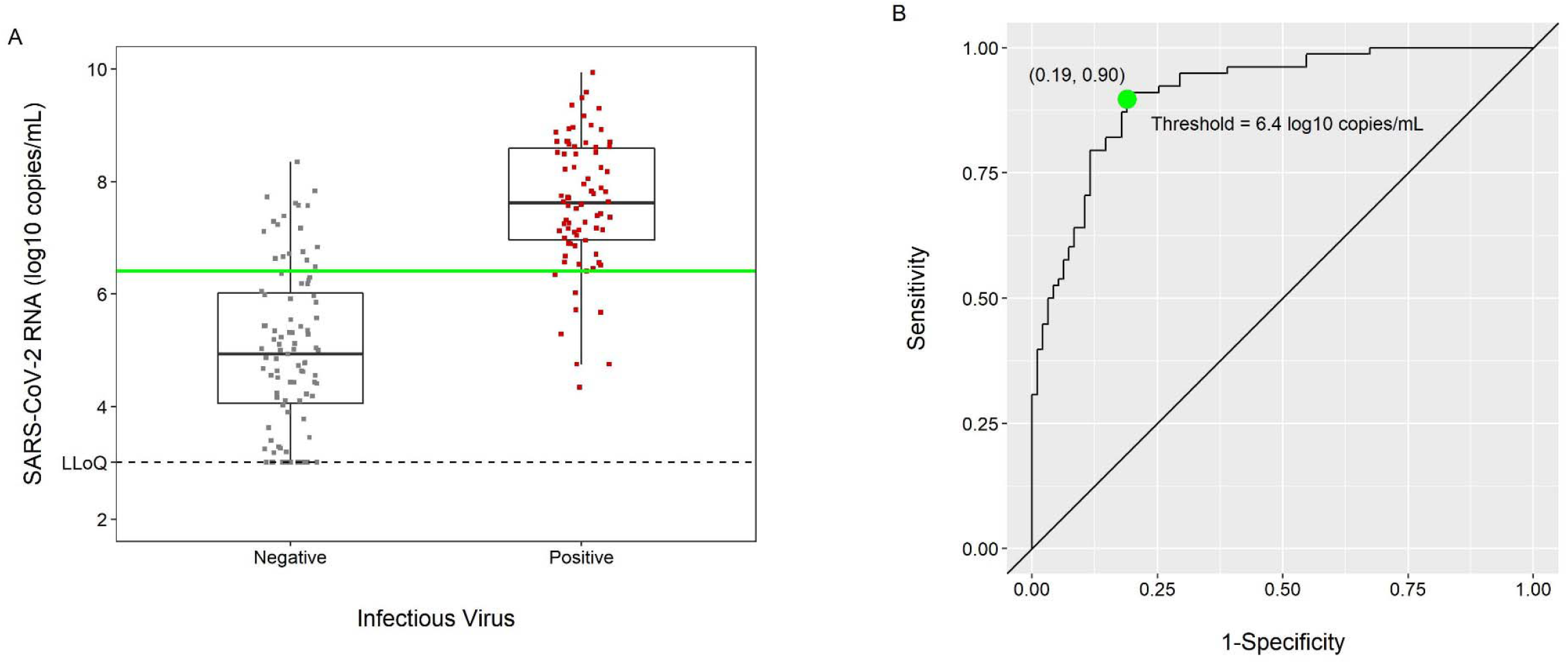
SARS-CoV-2 viral RNA by infectious virus culture – ROC Curve. Panel A shows boxplots of viral RNA measured via qRT-PCR displayed by infectious virus status, with culture negative in grey and culture positive in red; each dot represents a participant. Panel B shows a receiver operator characteristic (ROC) curve. A threshold of 6.4 viral RNA log_10_ copies/mL maximized sensitivity and specificity for SARS-CoV-2 infectious virus isolation and is represented by the green line on the boxplots and the corresponding green dot on the ROC curve.

**Supplementary Figure 4.**
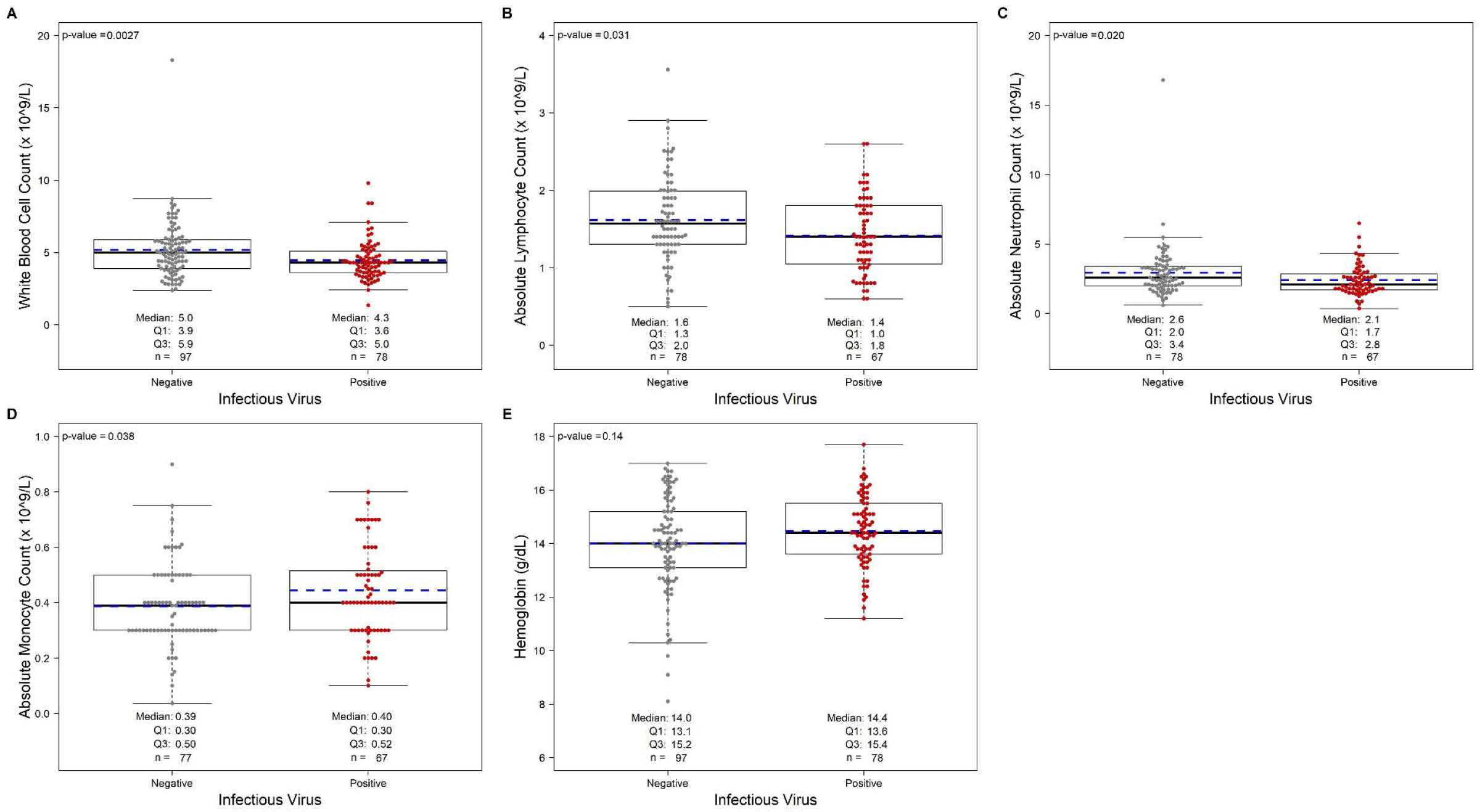
Associations between hematology measures and infectious SARS-CoV-2 virus. Panels A-E: Five hematology measures are displayed by infectious virus status, with infectious virus culture negative shown in grey and culture positive shown in red; each dot represents a participant. Solid lines on boxplots display the median and 25th-75th percentile (mean = blue dashed line).

**Supplementary Figure 5.**
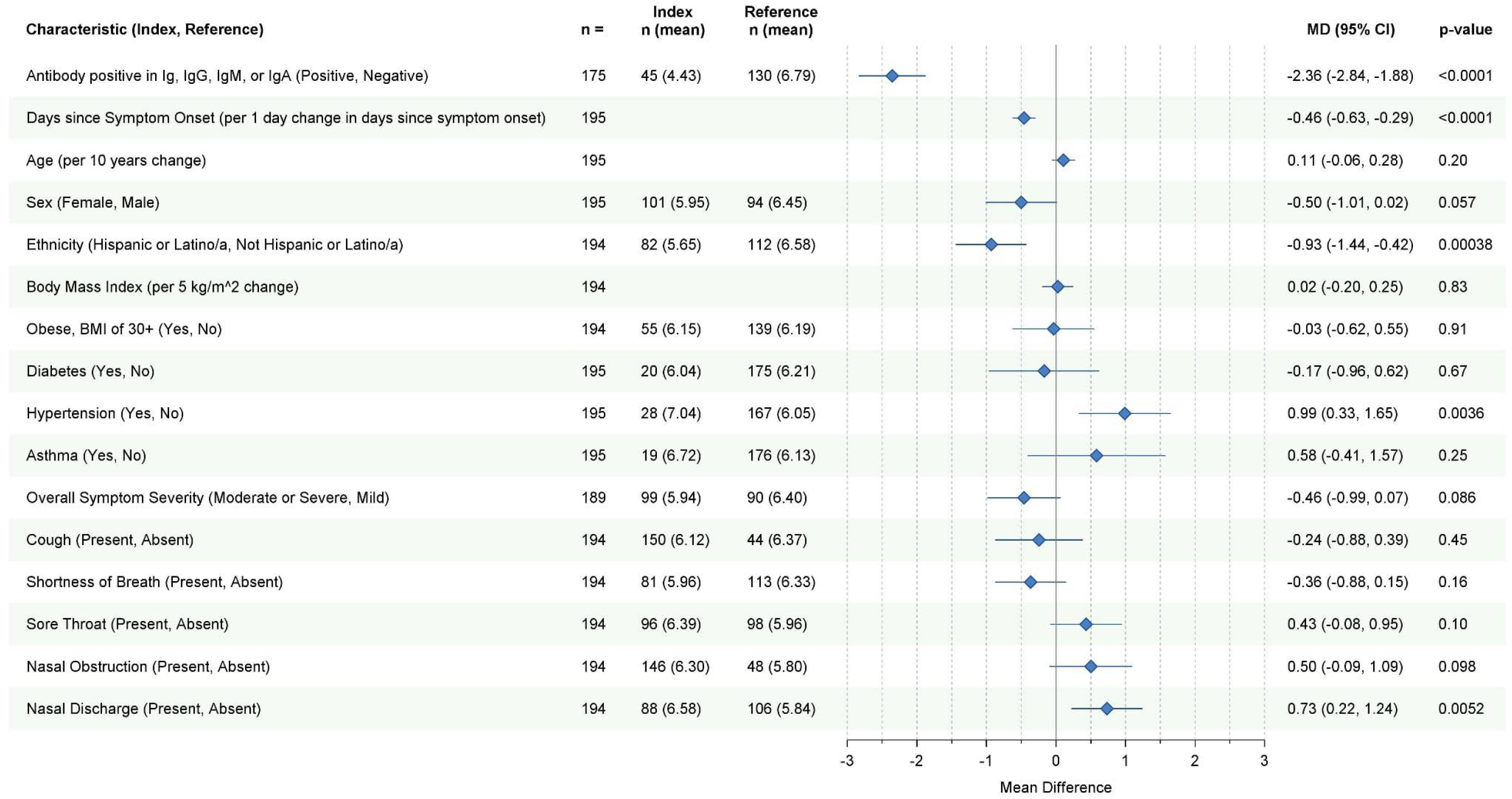
Host factor associations with SARS-CoV-2 viral RNA level. Bivariate analyses are shown. Mean differences in SARS-CoV-2 viral RNA (log_10_ copies/mL) measured via qRT-PCR were estimated with a corresponding 95% confidence interval. MD = mean difference.

